# Radiographic evaluation of lumbar intervertebral disc height index: an intra and inter-rater agreement and reliability study

**DOI:** 10.1101/2021.11.29.21267030

**Authors:** Xiaolong Chen, Stone Sima, Harvinder S. Sandhu, Jeff Kuan, Ashish D. Diwan

## Abstract

**Purpose:** To evaluate intra- and inter-rater agreement and reliability of seven reported disc height index (DHI) measurement methods on standing lateral X-ray of lumbar spine.

**Methods:** The adult patients who had standing lateral X-ray of lumbar spine were recruited. Seven methods were used to measure DHI of each lumbar intervertebral disc level. Bland and Altman’s Limits of Agreement (LOA) with standard difference were calculated to examine intra- and inter-rater agreements between two out of seven methods for DHI. Intra-class correlations (ICC) with 95% confidence intervals were calculated to assess intra- and inter-rater reliability.

**Results:** The intra-rater reliability in DHI measurements for 288 participants were ICCs from 0.807 (0.794, 0.812) to 0.922 (0.913, 0.946) by rater 1 (SS) and from 0.827 (0.802, 0.841) to 0.918 (0.806, 0.823) by rater 2 (XC). Method 2, 3, and 5 on all segmental levels had bias (95% CI does not include zero) or/and out of the acceptable cut-off proportion (>50%). A total of 609 outliers in 9174 segmental levels’ LOA range. Inter-rater reliability was good-to-excellent in all but method 2 (0.736 (0.712, 0.759)) and method 5 (0.634 (0.598, 0.667)). ICCs of related lines to good-to-excellent reliability methods was excellent in all but only indirect line in method 1 and 4 (ICCs lie in the range from 0.8 to 0.9).

**Conclusion:** Following structured protocol, intra- and inter-rater reliability was good-to-excellent for most DHI measurement methods on X-ray. However, in the presence of vertebral rotation, one should exercise caution in using complicated methods to define vertebral landmarks.

## Introduction

Low back pain (LBP) is the leading cause of disability worldwide with a lifetime prevalence that exceeds 90% [1]. Within the vast differential of LBP, the degeneration of intervertebral disc (IVD) is considered as a significant contributor [2]. Radiological examinations of the morphologic characteristic of lumbar IVD such as height has been found to be related to the degeneration of IVD [3]. The change of IVD height influences the load-carrying capacity of the spinal column, and morphologic abnormalities such as IVD space narrowing and thinning have been potentially associated with acute or chronic disabilities of the lumbar spine [4]. However, there is a paucity of information using different methods to estimate the disc height (DH) and its clinical significance. Therefore, an accurate and efficient measurement for IVD height is required.

Compared with lying supine during the magnetic resonance imaging (MRI) and computed tomography (CT) scan, the standing X-ray of lumbar spine can better present the state of IVD under load. Therefore, X-ray is considered as the most frequently used technique despite known difficulties, both in interpretation and clinical significance of findings. Clinicians often rely on their own subjective interpretation of lumbar spine radiographs, however, numerous methods for DH using X-ray published in the literature have been described as more accurate, albeit, and more time consuming [5-10]. DH can be measured as an absolute value, although this may be influenced by the magnification and position of the patient on the scan. Simple values can be used in daily practice for quick comparisons. For more in-depth studies and more accurate readings, the disc height index (DHI) has been introduced. By normalising images, variations in the size of the vertebral column and position of the patient do not affect the final measurement and allow for a reliable analysis. Many DHI measurement methods of IVD has been discussed previously in the literature [5-10]. However, this lack of consensus leads to great inter- and even intra-rater variability. A simple and reproducible method to measure DHI is required.

Bland and Altman’s Limits of Agreement (LOA) is the most popular [11], and recommended statistical method for evaluation of agreement [12, 13]. The standard error of measurement (SEM) is similarly regarded as a suitable parameter of agreement, but is, however, sensitive to variability in the population [14]. Although recent study reported use of LOA for evaluating agreement of measurements on intervertebral disc morphology using MRI images [15], it is rarely used when evaluating agreement in the different measurements of DHI using X-ray. Therefore, we need to use LOA to evaluate intra- and inter-rater agreement and reliability of DHI using the previously reported methods [5-10].

The primary objective of this study is to evaluate intra- and inter-rater agreement and reliability of seven previously reported DHI measurement methods.

## Methods

Ethical approval was obtained from the Human Research Ethics Committee of the University of New South Wales (NRR-HC180423) for the intra- and inter-rater agreement and reliability study using repeated measurement methods of individuals’ X-rays.

### Design and Patients

The study is conducted as a retrospective review of radiological images, radiology reports, and demographic data of patients over the age of 18 years who had routine standing lateral X-ray of lumbar spine from St George MRI in Sydney (Australia) from March 2017 onwards. Only patients who signed the consent form to allow use of their de-identified data for research and auditing purposes were included in the present study. The patients who had a history of spine surgery were excluded from the study.

### Measurements

The standard standing lateral X-ray images of lumbar spine were assessed. The patient is naturally standing up, looking horizontally, hands resting on a vertical support, upper limbs relaxed, elbows half bent [16]. The corresponding radiology reports were read by the first author (XC). Seven methods were used to measure the DHI of each lumbar IVD level on standing later X-ray images (L1-L2, L2-L3, L3-L4, L4-L5, and L5-S1) [5-10]. The protocol and details of DHI measurement methods are presented as follows and showed in Electronic Supplementary Material 1 (ESM_1) and Fig. 1.

**Fig. 1.**
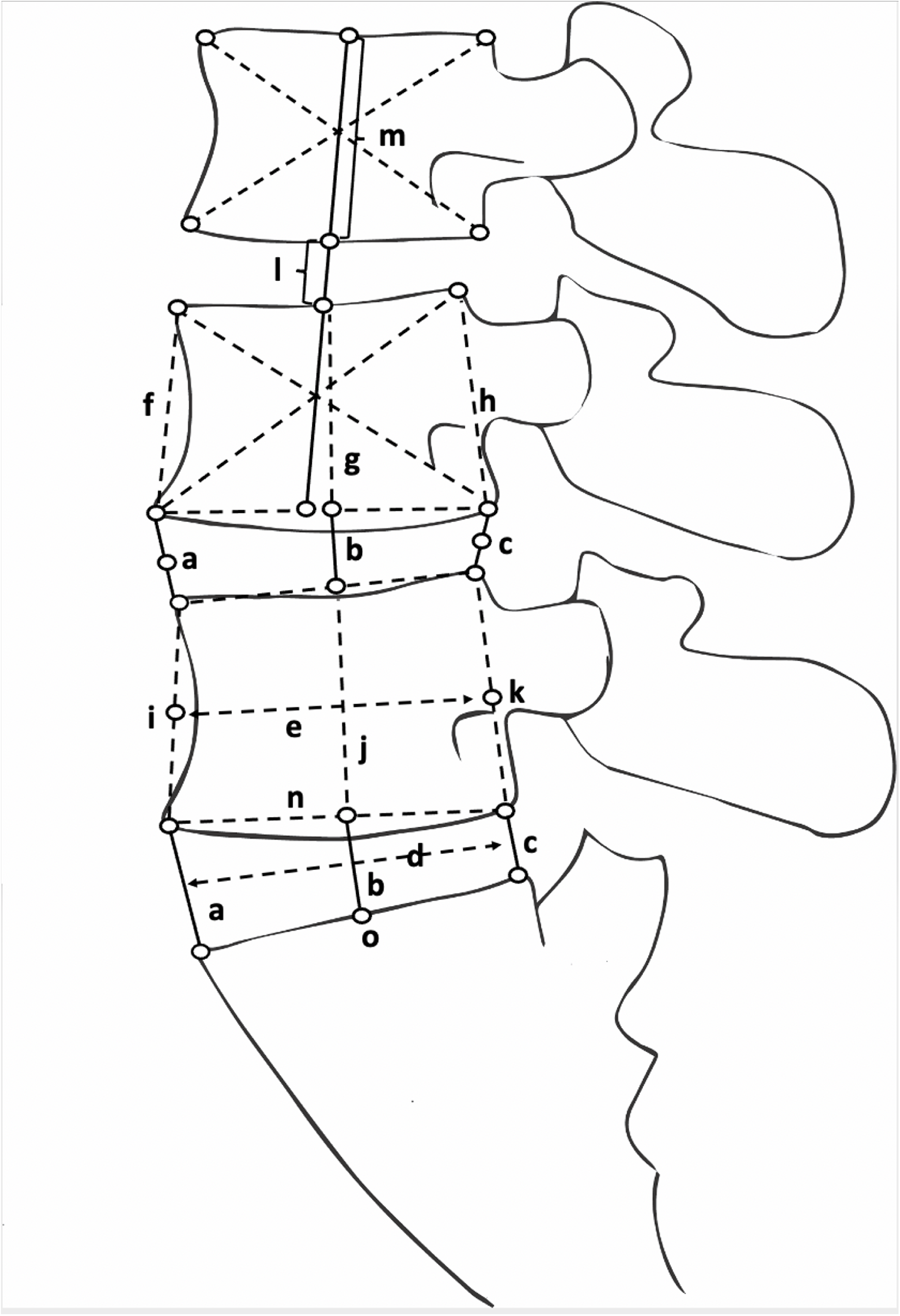
The details of disc height index (DHI) measurements Note: a: The shortest distance between the anterior edges of the neighbouring endplate will be recorded as the anterior disc height; b: The mid-disc height between the upper and lower bisection points is measured at the midpoint of vertebrae; c: The shortest distance between the posterior edges of the neighbouring endplate will be recorded as the posterior disc height; d: The disc diameter will be measured between the midpoints of the lines drawn from the endpoints of the superior vertebral endplate to the inferior; e: The sagittal diameter of the vertebral body from the anterior to posterior margin will be measured at the mid-vertebral level; f-h: The proximal vertebral body height will be measured from the anterior (f), middle (g), and posterior (h) portions of each respective disc level; i-k: The distal (DV) vertebral body height will be measured from the anterior (i), middle (j), and posterior (k) portions of each respective disc level; l, m: The mid-vertebral line is the line connecting the L3 and L4 centres. The centre of the vertebral body is a crossing point of 2 diagonal lines of each vertebral body (l is intervertebral disc height, m is intervertebral height); n: superior disc depth; o: inferior disc depth. According to the classification of related lines, line a, c, f, h, i, k, n and o are defined as direct lines and line b, d, e, g, j, l, and m are defined as indirect lines. Method 1: DHI = [(a+c)/d] *100% Method 2: DHI = (b/g) *100% or (b/j) *100% Method 3: DHI = (b/d) *100% Method 4: DHI = [(a+b+c)/3/e] *100% Method 5: DHI = (l/m) *100% Method 6: DHI = [2*(a+b+c)/((f+g+h)+(i+j+k))] *100% Method 7: DHI = [(a+c)/(n+o)] *100%

Method 1 of DHI is expressed as a ratio of the sum of anterior and posterior IVD height to disc diameter [5]. Method 2 of DHI is expressed as a ratio of the mid-disc height to mid-vertebral body height [6]. Method 3 of DHI is expressed as a ratio of the mid-disc height to disc diameter [6]. Method 4 of DHI is expressed as a ratio of the mean of anterior, middle, and posterior IVD height to the sagittal diameter of the proximal vertebral body [7]. Method 5 of DHI is expressed as a ratio of IVD height to vertebral height which cross the centre of adjacent vertebral bodies [8]. Method 6 of DHI is expressed as a ratio of the mean of anterior, middle, and posterior IVD height to the mean of proximal and distal vertebral body height [9]. Method 7 of DHI is expressed as a ratio of the sum of anterior and posterior IVD height to the sum of superior and inferior disc depth [10].

A quadrilateral was drawn to define the vertebral corners and minimize the affection of osteophytes. A line was drawn cross the potential points of each corner which was caused by the vertebral rotation for inexact body position during the scan and the anatomy deformity (such as scoliosis, vertebral rotation, and vertebral fracture). Mid-point of the line was identified as the real vertebral corner. Direct line was draw cross the two points which were located at the vertebral body. Indirect line was drawn cross the potential points which were location at direct lines.

If MRI scans already performed and presented in St Georgy MRI, the images were assessed the IVD degeneration. IVD degeneration is defined as the presence of at least one of the following: nucleus pulposus degeneration, IVD bulge or IVD herniation, annular tear, Modic changes of endplate, and Schmorl’s node [17-21]. Nucleus pulposus degeneration is defined as Pfirrmann grade ≥ 3 [22]. Participants were allocated into different groups (degeneration group and no degeneration group) based on the IVD degeneration status.

In order to reduce the potential bias due to difference of equipment and software, raters used Apple MacBook with integrated touchpads and the *InteleViewer*^*TM*^ diagnostic imaging software for measurement.

### Training and Blinding of Raters

Two raters conducted the measurements: one is a medical student (SS) who has no prior training in the interpretation of radiological images (Rater 1); the other is an experienced spine surgeon and back pain researcher (XC) with extensive experience in interpreting radiological images (Rater 2). Thirty participants from the final data collection period were randomly selected for training. Each rater reviewed the 30 cases independently, after which the cases were collectively reviewed, and consensus were reached on the measurement procedures. Once the raters reached an agreement on the measurement procedures, the data of these 30 cases was used to analysis the intra-rater reliability. The intra- and inter-rater agreement were tested between two out of seven measurements performed by each rater. The inter-rater reliability was tested between two raters who were purposely chosen to represent an inexperienced, and an experienced interpreter of radiological images.

To enhance the quality and applicability of the study, both raters were blinded in several aspects. Each rater was blinded to his own prior measurements and the findings of the other ratter. The order of participants was randomly changed between the two intra-rater measurement sessions. There was a 2-week interval between the first and second measurement sessions to lessen the likelihood of recognition of participants.

### Statistical analysis

Numeric variables are presented as mean ± standard deviation (SD). Categorical variables are summarized using counts (n) and percentages (%). The intra- and inter-rater agreement between two out of seven methods for DHI were analysed using Bland and Altman’s LOA. LOA is based on graphical techniques and provides a plot of mean differences (MDs) between the two methods of measurement (the bias), as well as the SD of the differences (Fig. 2). The 95% confident intervals (95% CI) of MDs were reported to describe the precision of the bias. If the 95% CI doesn’t include zero, it can be assumed that there is a bias. Furthermore, LOA was presented as a proportion of mean values for each method. The proportion will be calculated as follows: ((upper LOA +(−1*(lower LOA)))/(the mean)) *100%. Following previously published data, we consider percentages lower than 50% as an indicator of acceptable precision [15].

**Fig. 2.**
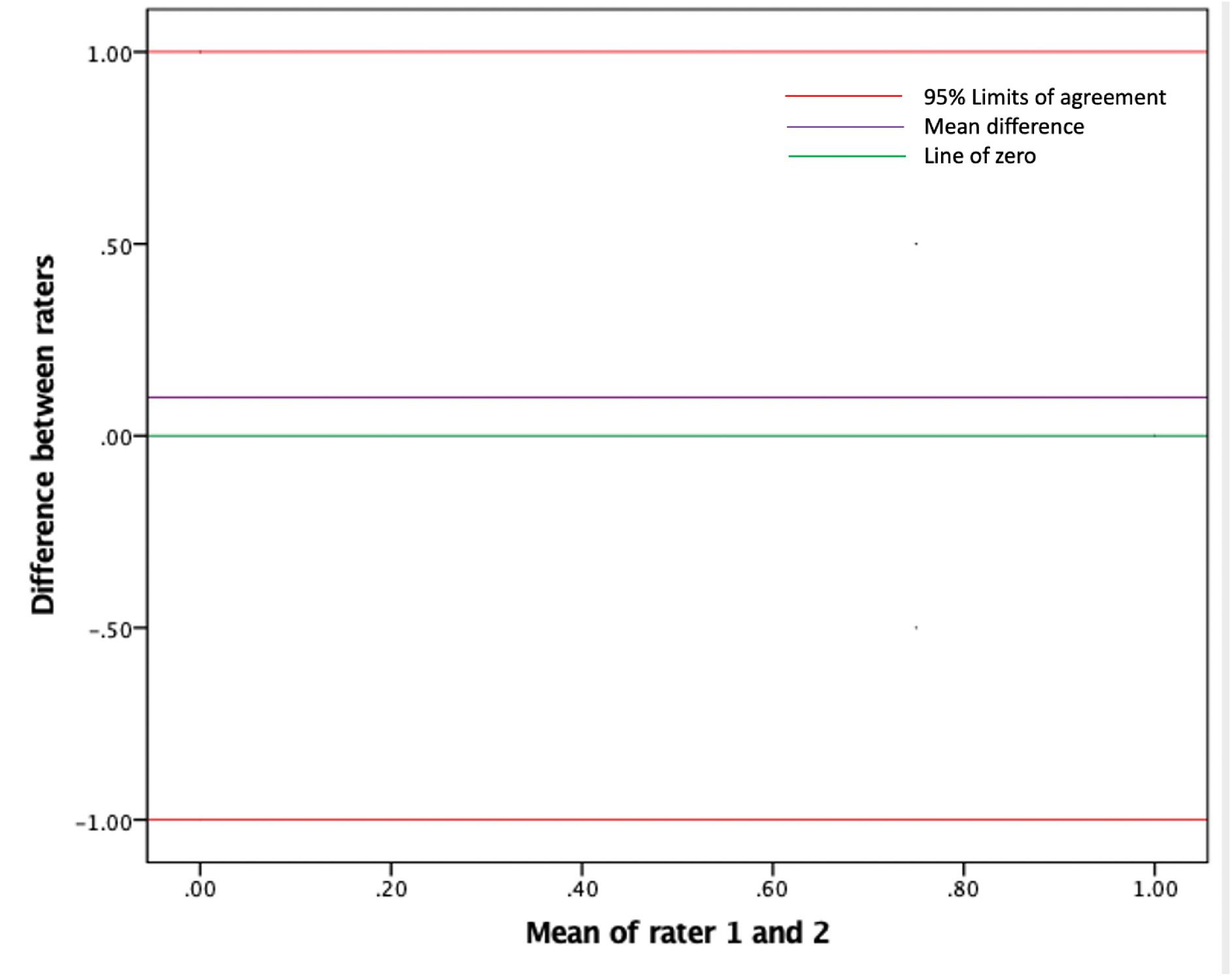
The Bland and Altman plot of Limits of Agreement (LOA) between two raters on the different measurements for disc height index (DHI). The y-axis shows the mean difference between raters’ measurements, and the x-axis shows the mean value of both raters’ measurements. The green line shows the range of mean difference includes zero. The purple line shows the mean difference between measurements. Red lines show the 95% LOA.

Intra-class correlation coefficient (ICC) estimates, and their 95% CI were calculated using SPSS statistical package version 24 (SPSS Inc, Chicago, IL) based on a single-rating, absolute-agreement, 2-way fixed-effects model for intra-rater reliability. Results of inter-rater reliability was evaluated with ICC based on a single-rating, consistency, 2-way random-effects model in all participants and different degeneration groups. Values of ICC less than 0.5, between 0.5 and 0.75, between 0.75 and 0.9, and greater than 0.90 are indicative of poor, moderate, good, and excellent reliability, respectively [23]. Subgroup analysis was performed based on different segmental level, the status of IVD degeneration, and different related lines (direct and indirect line).

### Factors analysis on the Bland and Altman’s plot

Potential factors for the data that were far above or below the LOA on the graphs were assessed and reported in a narrative form.

## Results

In total, the standing lumbar X-ray from 288 participants were included in this study for evaluation of both intra- and inter-rater reliability and agreement. There were 122 females and 166 males, all aged between 19 and 89 years. Of 367 lumbar levels with IVD degeneration in 278 participants who performed MRI scans (Table 1).

**Table 1.** Patient demographic and clinic-radiological information

| Parameter | Number of patients |
| --- | --- |
| F:M | 122:166 |
| Age | 47.67±16.79 |
| Diagnosis |  |
| Spondylolisthesis | 32 (11.1%) |
| Disc herniation | 57 (19.8%) |
| Spinal stenosis | 174 (60.4%) |
| Scoliosis | 11 (3.8%) |
| Normal | 88 (30.6%) |
| MRI scans (number of patients) | 278 (96.5%) |
| Intervertebral disc degeneration (number of patients) | 231 (83.1%) |
| Lumbar levels with intervertebral disc degeneration (total) | 367 |
| L1-L2 | 2 (0.5%) |
| L2-L3 | 0 |
| L3-L4 | 36 (9.8%) |
| L4-L5 | 160 (43.6%) |
| L5-S1 | 169 (46%) |

F: female; M: male; MRI: Magnetic resonance imaging

### Intra-rater reliability

The intra-rater reliability for DHI of all measurement methods, using ICC, was good-to-excellent from 0.807 (0.794, 0.812) to 0.922 (0.913, 0.946) by rater 1 and from 0.827 (0.802, 0.841) to 0.918 (0.806, 0.823) by rater 2, respectively (Table 2).

**Table 2.** Intra-rater measures’ reliability results

| Measurement method |  | N | Rater 1_ICC (95% CI) | Rater 2_ICC (95% CI) |
| --- | --- | --- | --- | --- |
| Method 1 | L1-L2 | 30 | 0.907 (0.902, 0.921) | 0.917 (0.908, 0.924) |
|  | L2-L3 | 30 | 0.867 (0.860, 0.882) | 0.866 (0.854, 0.884) |
|  | L3-L4 | 30 | 0.876 (0.861, 0.893) | 0.888 (0.876, 0.893) |
|  | L4-L5 | 30 | 0.822 (0.811, 0.843) | 0.858 (0.836, 0.873) |
|  | L5-S1 | 30 | 0.855 (0.841, 0.873) | 0.878 (0.856, 0.893) |
|  | All | 150 | 0.875 (0.872, 0.889) | 0.907 (0.902, 0.928) |
| Method 2 | L1-L2 | 30 | 0.821 (0.817, 0.842) | 0.845 (0.822, 0.864) |
|  | L2-L3 | 30 | 0.807 (0.794, 0.812) | 0.827 (0.802, 0.841) |
|  | L3-L4 | 30 | 0.922 (0.913, 0.946) | 0.912 (0.895, 0.946) |
|  | L4-L5 | 30 | 0.823 (0.812, 0.844) | 0.834 (0.828, 0.862) |
|  | L5-S1 | 30 | 0.842 (0.817, 0.881) | 0.868 (0.860, 0.890) |
|  | All | 150 | 0.848 (0.807, 0.855) | 0.871 (0.869, 0.889) |
| Method 3 | L1-L2 | 30 | 0.822 (0.810, 0.864) | 0.842 (0.838, 0.876) |
|  | L2-L3 | 30 | 0.843 (0.831, 0.861) | 0.861 (0.849, 0.873) |
|  | L3-L4 | 30 | 0.830 (0.815, 0.856) | 0.833 (0.805, 0.856) |
|  | L4-L5 | 30 | 0.853 (0.842, 0.883) | 0.869 (0.857, 0.883) |
|  | L5-S1 | 30 | 0.851 (0.812, 0.865) | 0.863 (0.841, 0.876) |
|  | All | 150 | 0.850 (0.832, 0.864) | 0.865 (0.844, 0.875) |
| Method 4 | L1-L2 | 30 | 0.882 (0.962, 0.896) | 0.892 (0.882, 0.898) |
|  | L2-L3 | 30 | 0.831 (0.822, 0.848) | 0.842 (0.812, 0.855) |
|  | L3-L4 | 30 | 0.877 (0.872, 0.882) | 0.879 (0.868, 0.885) |
|  | L4-L5 | 30 | 0.852 (0.834, 0.881) | 0.872 (0.865, 0.889) |
|  | L5-S1 | 30 | 0.916 (0.901, 0.923) | 0.918 (0.806, 0.823) |
|  | All | 150 | 0.879 (0.869, 0.912) | 0.887 (0.875, 0.914) |
| Method 5 | L1-L2 | 30 | 0.817 (0.801, 0.841) | 0.841 (0.823, 0.866) |
|  | L2-L3 | 30 | 0.854 (0.833, 0.867) | 0.858 (0.843, 0.872) |
|  | L3-L4 | 30 | 0.845 (0.840, 0.869) | 0.876 (0.855, 0.883) |
|  | L4-L5 | 30 | 0.861 (0.832, 0.877) | 0.873 (0.847, 0.879) |
|  | All | 120 | 0.858 (0.836, 0.873) | 0.871 (0.845, 0.881) |
| Method 6 | L1-L2 | 30 | 0.878 (0.848, 0.881) | 0.882 (0.878, 0.891) |
|  | L2-L3 | 30 | 0.822 (0.817, 0.881) | 0.871 (0.862, 0.889) |
|  | L3-L4 | 30 | 0.852 (0.827, 0.881) | 0.866 (0.854, 0.887) |
|  | L4-L5 | 30 | 0.856 (0.843, 0.883) | 0.884 (0.872, 0.894) |
|  | All | 120 | 0.859 (0.846, 0.879) | 0.878 (0.873, 0.888) |
| Method 7 | L1-L2 | 30 | 0.860 (0.842, 0.866) | 0.869 (0.851, 0.878) |
|  | L2-L3 | 30 | 0.851 (0.812, 0.865) | 0.863 (0.841, 0.876) |
|  | L3-L4 | 30 | 0.845 (0.814, 0.868) | 0.871 (0.855, 0.886) |
|  | L4-L5 | 30 | 0.855 (0.843, 0.865) | 0.865 (0.844, 0.875) |
|  | L5-S1 | 30 | 0.912 (0.878, 0.922) | 0.864 (0.848, 0.872) |
|  | All | 150 | 0.866 (0.844, 0.916) | 0.868 (0.858, 0.876) |

N: number of levels; ICC: Intraclass correlation coefficient; 95% CI: 95% confidence interval.

### Inter-rater agreement

#### Method 1

The 95% CI of mean difference of DHI on segmental level L3-L4 ranged between -0.006 and 0.005, with LOA ranging between -0.10 and 0.10 (LOA as proportion of mean values is 33.9%). The 95% CI of mean difference of DHI on segmental level L5-S1 ranged between -0.014 and 0.002, with LOA ranging between -0.15 and 0.13 (LOA as proportion of mean values is 46.7%) (Table 3 and Fig. 3A).

**Table 3.** Inter-rater measures agreement results

|  | Level | N | Mean | SD | Mean difference<br>(95% CI) | 95% LOA | LOA as proportion<br>of mean values (%) |
| --- | --- | --- | --- | --- | --- | --- | --- |
| **Method 1** | L1-L2 | 288 | 0.55 | 0.07 | 0.00 (-0.008, 0.008) | -0.14, 0.14 | 50.9 |
|  | L2-L3 | 288 | 0.55 | 0.05 | 0.01 (0.005, 0.016) * | -0.09, 0.11 | 36.4 |
|  | L3-L4 | 288 | 0.59 | 0.05 | 0.00 (-0.006, 0.005) | -0.10, 0.10 | 33.9 |
|  | L4-L5 | 288 | 0.61 | 0.09 | 0.01 (0.001, 0.022) * | -0.17, 0.19 | 59.0 |
|  | L5-S1 | 288 | 0.6 | 0.07 | -0.01 (-0.014, 0.002) | -0.15, 0.13 | 46.7 |
|  | All | 1440 | 0.58 | 0.07 | 0.00 (0.002, 0.007) * | -0.14, 0.14 | 48.3 |
| **Method 2** | L1-L2 | 288 | 0.36 | 0.07 | -0.03 (-0.034, -0.019) * | -0.17, 0.11 | 77.8 |
|  | L2-L3 | 288 | 0.38 | 0.07 | -0.03 (-0.039, -0.021) * | -0.17, 0.11 | 73.7 |
|  | L3-L4 | 288 | 0.39 | 0.07 | -0.03 (-0.036, -0.020) * | -0.17, 0.11 | 71.8 |
|  | L4-L5 | 288 | 0.39 | 0.06 | -0.02 (-0.026, -0.014) * | -0.14, 0.10 | 61.5 |
|  | L5-S1 | 288 | 0.37 | 0.06 | -0.02 (-0.024, -0.009) * | -0.14, 0.10 | 64.9 |
|  | All | 1440 | 0.38 | 0.07 | -0.02 (-0.033, -0.018) * | -0.16, 0.12 | 73.7 |
| **Method 3** | L1-L2 | 288 | 0.29 | 0.04 | -0.01 (-0.015, -0.006) * | -0.09, 0.07 | 55.2 |
|  | L2-L3 | 288 | 0.3 | 0.04 | -0.01 (-0.019, -0.010) * | -0.09, 0.07 | 53.3 |
|  | L3-L4 | 288 | 0.32 | 0.04 | -0.01 (-0.014, -0.005) * | -0.09, 0.07 | 50.0 |
|  | L4-L5 | 288 | 0.3 | 0.03 | 0.00 (-0.009, -0.001) * | -0.06, 0.06 | 40.0 |
|  | L5-S1 | 288 | 0.27 | 0.04 | 0.00 (-0.003, 0.006) | -0.08, 0.08 | 59.3 |
|  | All | 1440 | 0.3 | 0.04 | -0.01 (-0.021, -0.009) * | -0.09, 0.07 | 53.3 |
| **Method 4** | L1-L2 | 288 | 0.29 | 0.03 | -0.01 (-0.013, -0.005) * | -0.07, 0.05 | 41.4 |
|  | L2-L3 | 288 | 0.9 | 0.09 | -0.02 (-0.033, -0.013) * | -0.20, 0.16 | 40.0 |
|  | L3-L4 | 288 | 0.31 | 0.03 | -0.01 (-0.012, -0.006) * | -0.07, 0.05 | 38.7 |
|  | L4-L5 | 288 | 0.31 | 0.03 | 0.00 (-0.008, 0.002) | -0.06, 0.06 | 38.7 |
|  | L5-S1 | 288 | 0.3 | 0.04 | -0.01 (-0.016, -0.008) * | -0.09, 0.07 | 53.3 |
|  | All | 1440 | 0.42 | 0.05 | -0.01 (-0.018, -0.008) * | -0.11, 0.09 | 47.6 |
| **Method 5** | L1-L2 | 288 | 0.36 | 0.13 | -0.02 (-0.037, -0.008) * | -0.27, 0.23 | 138.9 |
|  | L2-L3 | 288 | 0.37 | 0.08 | -0.02 (-0.030, -0.012) * | -0.18, 0.14 | 86.5 |
|  | L3-L4 | 288 | 0.37 | 0.06 | -0.01 (-0.021, -0.078) * | -0.13, 0.11 | 64.9 |
|  | L4-L5 | 288 | 0.36 | 0.05 | -0.01 (-0.012, 0.003) | -0.11, 0.09 | 55.6 |
|  | All | 1440 | 0.37 | 0.09 | -0.02 (-0.021, -0.008) * | -0.20, 0.16 | 97.3 |
| **Method 6** | L1-L2 | 288 | 0.34 | 0.04 | -0.01 (-0.012, -0.003) * | -0.10, 0.06 | 47.1 |
|  | L2-L3 | 288 | 0.35 | 0.04 | -0.01 (-0.013, -0.005) * | -0.10, 0.06 | 45.7 |
|  | L3-L4 | 288 | 1.41 | 0.06 | 0.00 (-0.003, 0.010) | -0.12, 0.12 | 17.0 |
|  | L4-L5 | 288 | 0.4 | 0.04 | 0.00 (-0.009, 0.001) | -0.08, 0.08 | 40.0 |
|  | All | 1440 | 0.62 | 0.04 | -0.00 (-0.010, 0.003) | -0.08, 0.08 | 25.8 |
| **Method 7** | L1-L2 | 288 | 0.27 | 0.04 | 0.00 (-0.002, 0.006) | -0.08, 0.08 | 59.3 |
|  | L2-L3 | 288 | 0.28 | 0.02 | 0.01 (0.003, 0.008) * | -0.03, 0.05 | 28.6 |
|  | L3-L4 | 288 | 0.29 | 0.02 | 0.00 (-0.003, 0.003) | -0.04, 0.04 | 27.6 |
|  | L4-L5 | 288 | 0.31 | 0.05 | 0.00 (0.001, 0.010) * | -0.10, 0.10 | 64.5 |
|  | L5-S1 | 288 | 0.3 | 0.03 | 0.00 (-0.006, 0.001) | -0.06, 0.06 | 40.0 |
|  | All | 1440 | 0.29 | 0.03 | 0.00 (0.002, 0.010) * | -0.06, 0.06 | 41.4 |

N: number of levels; SD: standard deviation; CI: confidential intervals; LOA: Limits of Agreement
\* Bias was considered present if the 95% CI did not include zero.

**Fig. 3.**
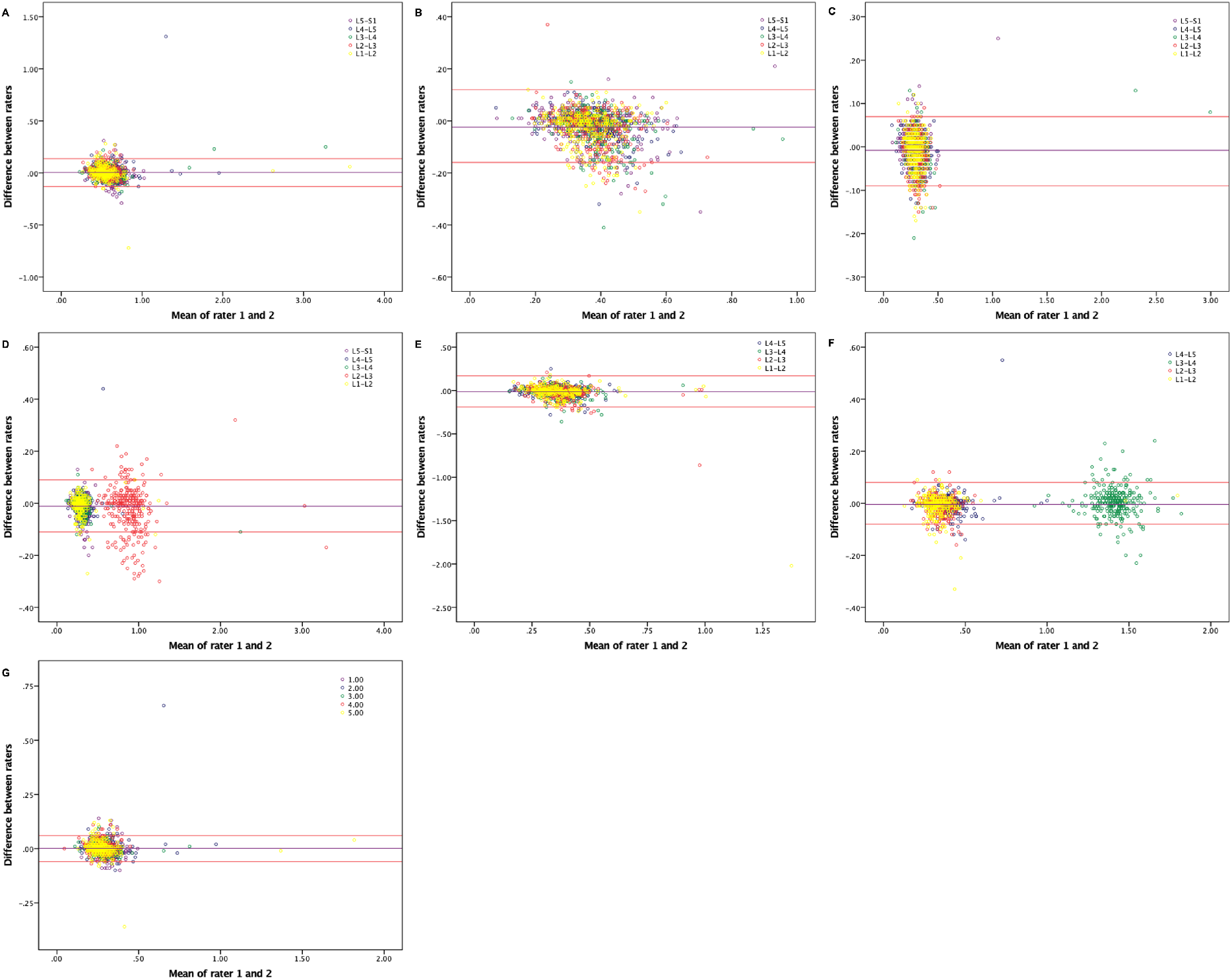
The Bland and Altman plot showing the relationship between mean values and differences between rater 1 and rater 2 on the measurements of DHI using two out of seven reported methods (A: method 1; B: method 2; C: method 3; D: method 4; E: method 5; F: method 6; G: method 7). Mean difference with 95% confidence intervals (CI) of the measurements between rater 1 and rater 2 was reported to describe the precision of the bias. The purple line shows the mean difference between measurements. Red lines show the 95% Limits of Agreement (LOA), between which 95% of all measurement differences are located.

#### Method 2, 3, and 5

The 95% CI of mean difference of DHI on all lumbar levels did not include zero or LOA as proportion of mean values is more than 50% (Table 3, Fig. 3B, Fig. 3C, and Fig. 3E).

#### Method 4

The 95% CI of mean difference of DHI on segmental level L4-L5 ranged between -0.008 and 0.002, with LOA ranging between -0.06 and 0.06 (LOA as proportion of mean values is 38.7%) (Table 3 and Fig. 3D).

#### Method 6

The 95% CI of mean difference of DHI on segmental level L3-L4 ranged between -0.003 and 0.010, with LOA ranging between -0.12 and 0.12 (LOA as proportion of mean values is 17%). The 95% CI of mean difference of DHI on segmental level L4-L5 ranged between -0.009 and 0.001, with LOA ranging between -0.08 and 0.08 (LOA as proportion of mean values is 40%) (Table 3 and Fig. 3F).

#### Method 7

The 95% CI of mean difference of DHI on segmental level L3-L4 ranged between -0.003 and 0.003, with LOA ranging between -0.04 and 0.04 (LOA as proportion of mean values is 27.6%). The 95% CI of mean difference of DHI on segmental level L5-S1 ranged between -0.006 and 0.001, with LOA ranging between -0.06 and 0.06 (LOA as proportion of mean values is 40%) (Table 3 and Fig. 3G).

### Inter-rater reliability

The inter-rater reliability for measurements of DHI was good-to-excellent in all but method 2 and 5 (ICCs ranged from 0.634 (0.598, 0.667) to 0.984 (0.982, 0.985); method 2: 0.736 (0.712, 0.759); method 5: 0.634 (0.598, 0.667)) (Table 4).

**Table 4.** Inter-rater measures’ reliability results

| Measurement | Level | N | Inter-rater_ICC (95% CI) |
| --- | --- | --- | --- |
| Method 1 | L1-L2 | 288 | 0.956 (0.945, 0.965) |
|  | L2-L3 | 288 | 0.877 (0.847, 0.901) |
|  | L3-L4 | 288 | 0.972 (0.964, 0.977) |
|  | L4-L5 | 288 | 0.844 (0.807, 0.874) |
|  | L5-S1 | 288 | 0.852 (0.817, 0.881) |
|  | All | 1440 | 0.927 (0.919, 0.934) |
| Method 2 | L1-L2 | 288 | 0.641 (0.568, 0.705) |
|  | L2-L3 | 288 | 0.620 (0.543, 0.686) |
|  | L3-L4 | 288 | 0.693 (0.628, 0.749) |
|  | L4-L5 | 288 | 0.779 (0.729, 0.821) |
|  | L5-S1 | 288 | 0.849 (0.813, 0.878) |
|  | All | 1440 | 0.736 (0.712, 0.759) |
| Method 3 | L1-L2 | 288 | 0.718 (0.657, 0.770) |
|  | L2-L3 | 288 | 0.781 (0.731, 0.822) |
|  | L3-L4 | 288 | 0.980 (0.975, 0.984) |
|  | L4-L5 | 288 | 0.866 (0.834, 0.892) |
|  | L5-S1 | 288 | 0.888 (0.860, 0.910) |
|  | All | 1440 | 0.936 (0.930, 0.942) |
| Method 4 | L1-L2 | 288 | 0.954 (0.942, 0.963) |
|  | L2-L3 | 288 | 0.938 (0.922, 0.950) |
|  | L3-L4 | 288 | 0.977 (0.972, 0.982) |
|  | L4-L5 | 288 | 0.872 (0.841, 0.897) |
|  | L5-S1 | 288 | 0.824 (0.783, 0.858) |
|  | All | 1440 | 0.984 (0.982, 0.985) |
| Method 5 | L1-L2 | 288 | 0.500 (0.409, 0.582) |
|  | L2-L3 | 288 | 0.726 (0.666, 0.776) |
|  | L3-L4 | 288 | 0.728 (0.669, 0.778) |
|  | L4-L5 | 288 | 0.761 (0.708, 0.805) |
|  | All | 1152 | 0.634 (0.598, 0.667) |
| Method 6 | L1-L2 | 288 | 0.951 (0.938, 0.961) |
|  | L2-L3 | 288 | 0.847 (0.811, 0.877) |
|  | L3-L4 | 288 | 0.862 (0.829, 0.889) |
|  | L4-L5 | 288 | 0.886 (0.859, 0.909) |
|  | All | 1152 | 0.995 (0.995, 0.996) |
| Method 7 | L1-L2 | 288 | 0.959 (0.949, 0.968) |
|  | L2-L3 | 288 | 0.891 (0.864, 0.913) |
|  | L3-L4 | 288 | 0.927 (0.909, 0.942) |
|  | L4-L5 | 288 | 0.840 (0.802, 0.871) |
|  | L5-S1 | 288 | 0.867 (0.835, 0.893) |
|  | All | 1440 | 0.916 (0.908, 0.924) |

N: number of levels; ICC: Intraclass correlation coefficient; 95% CI: 95% confidential intervals

#### Temporal analysis

Based on different segmental level, ICCs for DHI on segment level L1-L2 was moderate in method 2, 3, and 5 groups (ICC: 0.641 (0.568, 0.705), 0.718 (0.657, 0.770), 0.500 (0.409, 0.582)). ICCs for DHI on segment level L2-L3 was moderate in method 2 and 5 groups (ICC: 0.620 (0.543, 0.686), 0.726 (0.666, 0.776)). ICCs for DHI on segment level L3-L4 was moderate in method 2 and 5 groups (ICC: 0.693 (0.628, 0.749), 0.728 (0.669, 0.778)) (Table 4).

Based on the status of IVD degeneration, ICCs of DHI on all segmental levels in degeneration group and no degeneration group have a similar range based on the classification criterion for poor, moderate, good, and excellent reliability (ESM_2_Table 1).

ICCs of related lines to good-to-excellent reliability methods was excellent in all but only indirect line in method 1 and 4 (ICCs lie in the range from 0.8 to 0.9, ESM_2_Table 2).

#### Factors analysis on the Bland and Altman plot

A total of 609 outliers in 9174 segmental levels’ data includes 57 outliers in the method 1 group, 65 outliers in the method 2 group, 171 outliers in the method 3 group, 182 outliers in the method 4 group, 37 outliers in the method 5 group, 42 outliers in the method 6 group, and 55 outliers in the method 7 group (ESM_2_Table 3). The nucleus pulposus degeneration (394) and disc herniation (186) affected the raters to distinguish vertebral corners and structural boundaries.

## Discussion

The reduction of IVD height is the key point in the pathological process of IVD degeneration, and the diseases of lumbar degeneration often demonstrate the reduction of IVD height in the radiographic images. Therefore, a reproducible method to measure IVD height is required. To be the best of our knowledge, this is the first study to investigate the intra- and inter-rater reliability and agreement of previously reported DHI methods to measure DH on the standing lateral lumbar X-ray images. We used a structured protocol including descriptions of testing positions, standard training session of measurements on images for raters, unified measurement platform and tools, and blinding of raters [24].

Although the measurements on X-ray images would be affected by body posture and vertebral position of the patient on the scan and the experience of raters [6, 15, 25-27], our study still shown that intra-rater reliability was good-to-excellent for all the seven DHI assessment methods on X-ray images by both inexperienced and experienced raters. A possible explanation is the existence of division in the process of calculating the DHI, which can minimize the measurement bias by the inconsistent magnification and vertebral position on the X-ray scan. We posit the systematic training and structured protocol to conduct the measurement to be the other main cause of the good-to-excellent intra-rater reliability on DHI measurement methods on X-ray images. Therefore, the systematic training before measurement and a standard measurement process following structured protocol could provide a good-to-excellent intra-rater reliability for the DHI measurement on X-ray images.

Agreement is commonly used to evaluate how well the measurements produced by two raters, devices or systems agree with each other, while reliability is concerned with measurement error plus the variability between study objects and the focus is distinction between persons [15, 28]. Previously published study recommended reporting inter-rater agreement parameters via LOA, and further, when reporting reliability using ICC, they should be reported together with error estimates such as the standard error of the mean [28].

Following the results of Bland and Altman’s LOA, we found that the DHI measurements in method 2, 3, and 5 on all segmental levels and method 1, 4, 6 and 7 on some special segmental levels had bias or/and out of the acceptable cut-off proportion. Due to different numbers of indirect lines in each method, it indicates a poor-to-moderate consensus regarding the anatomical delineation on the length measurements between the two raters. These were consistent with the status of ours’ study that all indirect lines involved in method 2 and 5 and partial indirect lines involved in method 1, 3, and 4 with a poor-to-moderate agreement. Meanwhile, nucleus pulposus degeneration and disc herniation were showed to impact of the inter-rater agreement on distinguish vertebral corners and structural boundaries.

This study uses both LOA and reliability to express reproducibility. The inter-rater reliability was good-to-excellent in all but method 2 and 5. Although IVD degeneration can cause discs to lose height and might potentially affect the accuracy and agreement of DHI measurement [9], our findings denied the influence of IVD degeneration on the inter-rater reliability results in different measurement methods on DHI (ESM_2_Table 1). The potential risk factors for the moderate inter-rater reliability on DHI measurement in method 2 and 5 include measurement bias of indirect lines and other bias from anatomical structure. Due to use of multiple indirect lines in method 2 and 5, the potential secondary measurement bias following the first bias by the inexact positioning of vertebral corners and indistinguishable IVD boundaries between structures during drawing the direct line might cause the moderate inter-reliability. This indicates that a complicated measurement method would cause a poor-to-moderate consensus between raters. Despite good-to-excellent inter-rater reliability on DHI measurement in method 1, 3, 4, 6, and 7, there still showed relatively poor results of inter-rater reliability on indirect lines in method 1 and 4 (ESM_2_Table 2). Therefore, the accurate and effective determination of vertebral corners for direct line can significantly reduce the measurement error. As for the positioning of vertebral corners, two possible interfering factors could be the presence of osteophytes and the rotation of vertebral body, hence, modifying the visual appearance of the vertebra [6, 15, 29]. While our structured protocol could minimize the influence of osteophytes on marking the corners, it can’t provide a method to avoid the objective factor that leads to vertebral rotation. For instance, upper vertebral rotation by IVD no perpendicular to the projection might be the reason for moderate inter-rater reliability of DHI measurements on upper segmental level (L1-L2 and L2-L3). Meanwhile, the shorter DH of the upper IVD could induce cumulative error in the marking of vertebral corners, which was posited to be the other reason. We couldn’t find studies that definitively discussed any of these factors regarding similar problems with measurement bias of indirect lines, vertebral rotation, or boundary distinction. However, we still thought that these could be the main reasons why some ICCs of inter-rater reliability were moderate. As it stands, our study potentially showed that there was a good-to-excellent intra- and inter-rater reliability and agreement on the DHI measurements in method 7 for all IVD segmental levels. For future use of these methods, specification in advance of measurements, and persistent implementation of detailed protocol for the location of projection, measurement of indirect lines, and dealing with vertebral rotation, should be conducted by all raters.

### Study limitation and future study

Several methodological issues require consideration. First, the potential measurement error due to notably inexact definition of anatomic measurement points, the location of projection during the scan, definition of standard process to fix vertebral rotation, and intra- and inter-rater variation, despite a structural protocol being provided to raters. Future, a standardized protocol to assess DHI was required. Second, due to the difficulties in distinguishing the boundary of disc on X-ray, the raters can only use point-based measurement method instead of area-based method. Third, the acceptable precision of the range of LOA set at 50% following previously published data would affect the results [15]. Fourth, due to the different reference values of each DHI method, the direct comparison between two out of seven measurement methods can’t be done. Finally, the aim of this study was to establish reproducibility and reliability, not to report prevalence or reference values for either a general or a clinical population. Future multicenter study on the validity of different measurement methods is needed.

## Conclusion

The intra-rater and most inter-rater reliability for DHI measurement was good-to-excellent for different methods following a structured protocol. However, the inter-rater reliability was moderate in some DHI measurement methods, indicating difficulties in the performance of these tests. The complicated methods (more indirect lines) and IVD degeneration (nucleus pulposus degeneration and disc herniation) potentially affected the agreement on inter-rater measurements. Future multicenter study on the validity of different measurement methods following a standardized protocol is needed.

## Data Availability

All data produced in the present study are available upon reasonable request to the authors

## Acknowledgment

The authors would like to thank Vivek A.S. Ramakrishna (PhD Candidate, Spine Labs in St. George & Sutherland Clinical School and School of Mechanical and Manufacturing Engineering, University of New South Wales (UNSW)) for drawing the Figure 1 in this manuscript.

## ESM_1: The protocol of disc height index (DHI) measurement

Patients will have erect, standing lateral (LR) and anteroposterior (AP) X-rays taken at various time points (per study schema). Hard copies of X-rays will be printed and will also be stored as digital images from the *InteleViewer*^*TM*^ diagnostic imaging software/PACS system of the radiology provider. Care will be taken to maintain the aspect ratio of the digital images.

### (1) Standing Position of Patients

The patient is naturally standing up, looking horizontally, hands resting on a vertical support, upper limbs relaxed, elbows half bent.

The projection: The projection direction is perpendicular to the third vertebral body of the lumbar spine.

### (2) The Quality of Standing Lateral X-ray Images of Lumbar Spine

The entire lumbar spine should be visible from T12-L1 - L5-S1 (superior to include the T12-L1, inferior to include the sacrum, anterior to include the anterior border of the lumbar vertebral bodies, and posterior to include all elements of the posterior column, particularly the spinous processes). Superimposition of the greater sciatic notches, the superior articulating facets and the superior and inferior endplates. This indicates a true lateral has been achieved. Adequate image penetration and image contrast is evident by clear visualization of lumbar vertebral bodies, with both trabecular and cortical bone demonstrated.

### (3) Measurement Methods

The contour of the vertebral body and upper and lower endplate of adjacent vertebral will be indicated by lines. The vertebral corners could be located and confirmed.

A quadrilateral will be drawn to define the vertebral corners and minimize the affection of osteophytes.

A line is drawn cross the potential points of each corner which can be caused by the vertebral rotation for inexact body position during the scan and the anatomy deformity (such as scoliosis, vertebral rotation, and vertebral fracture).

Mid-point of the line will be identified as the real vertebral corner.

### (4) Potential Bias

In order to reduce the potential bias due to difference of equipment and software, Apple MacBook with integrated touchpads/Computer with Microsoft Windows and the *InteleViewer*^*TM*^ diagnostic imaging software/PACS system will be used for measurement.

**ESM_2_Table 1.**
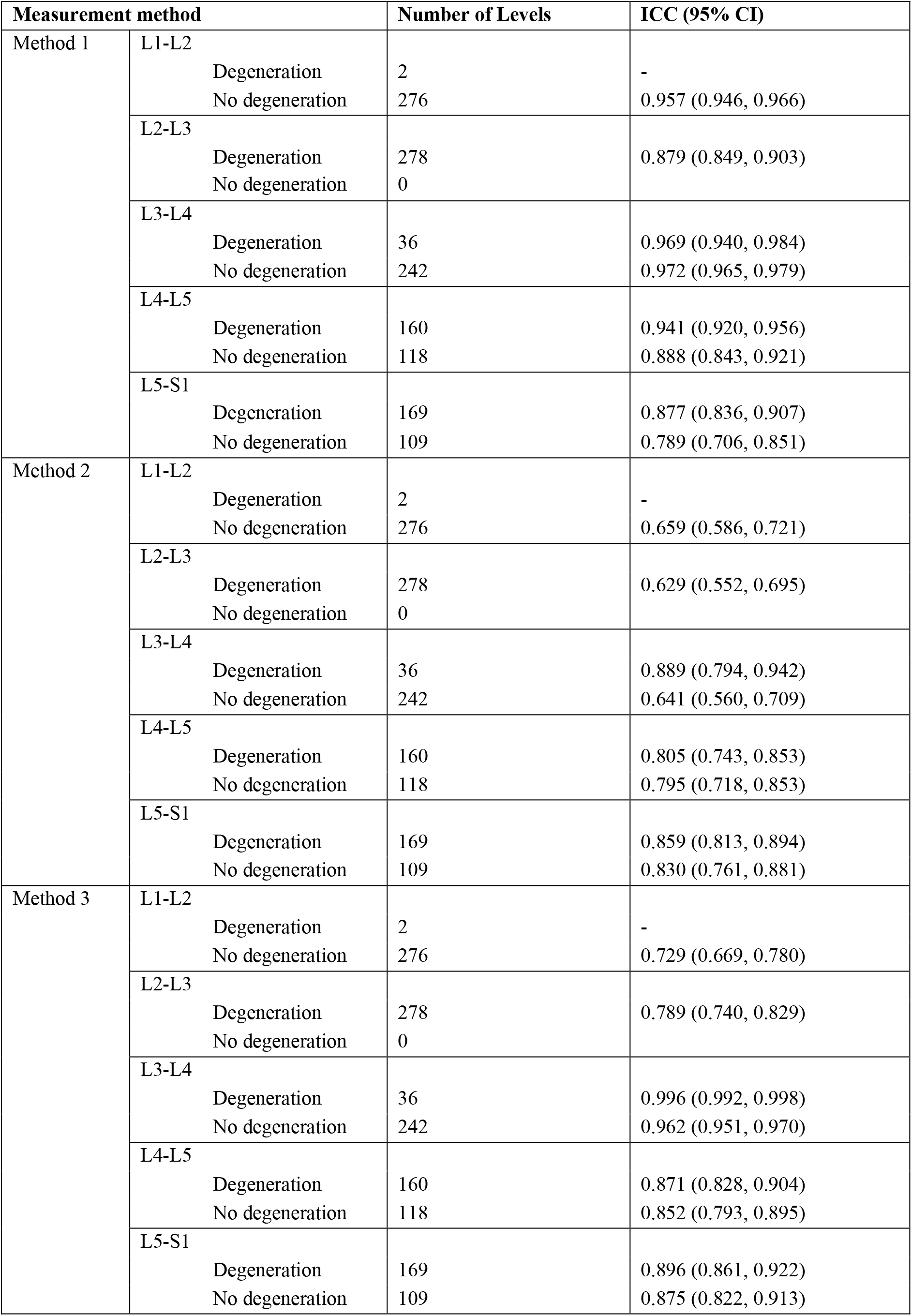
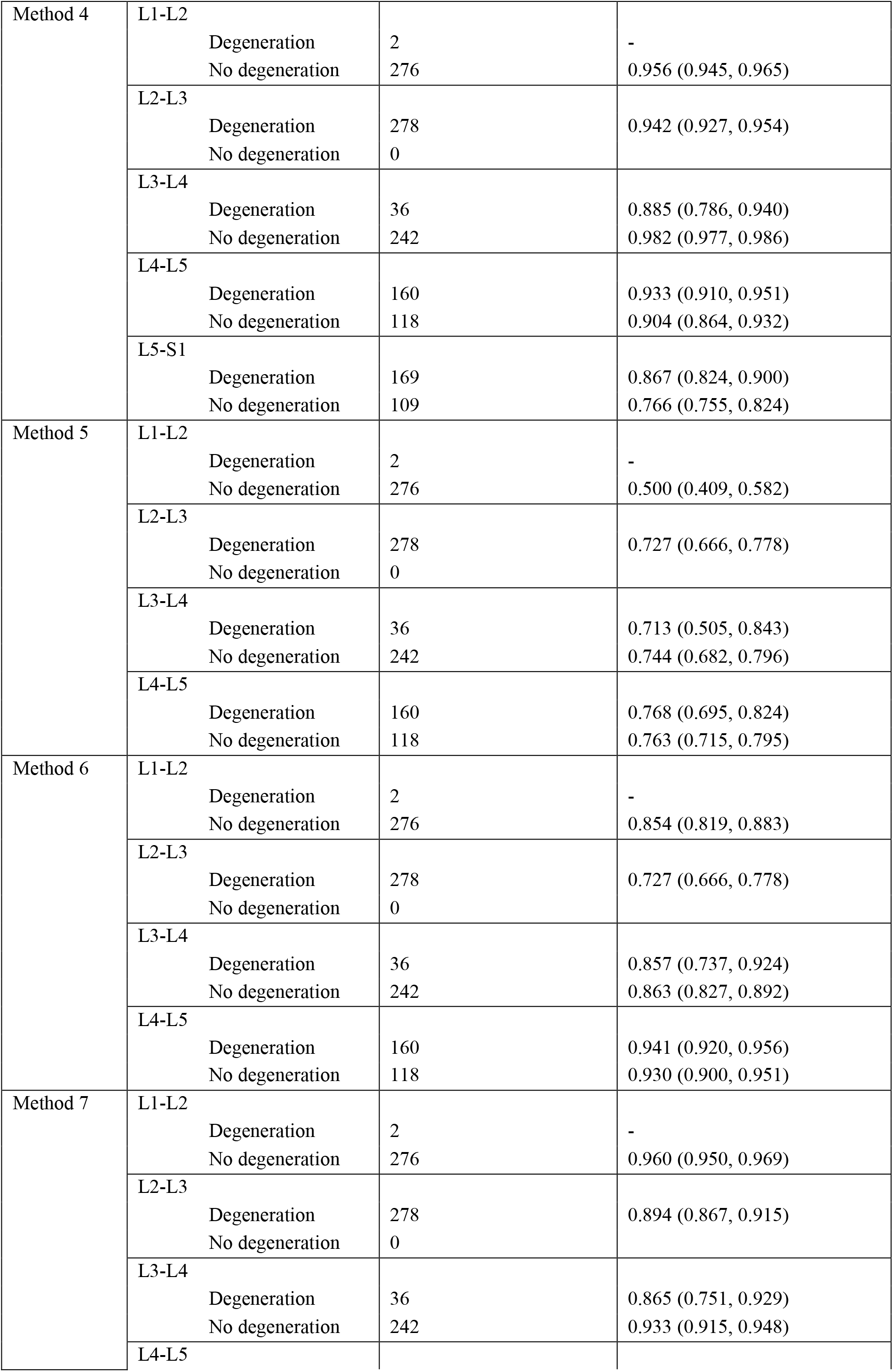
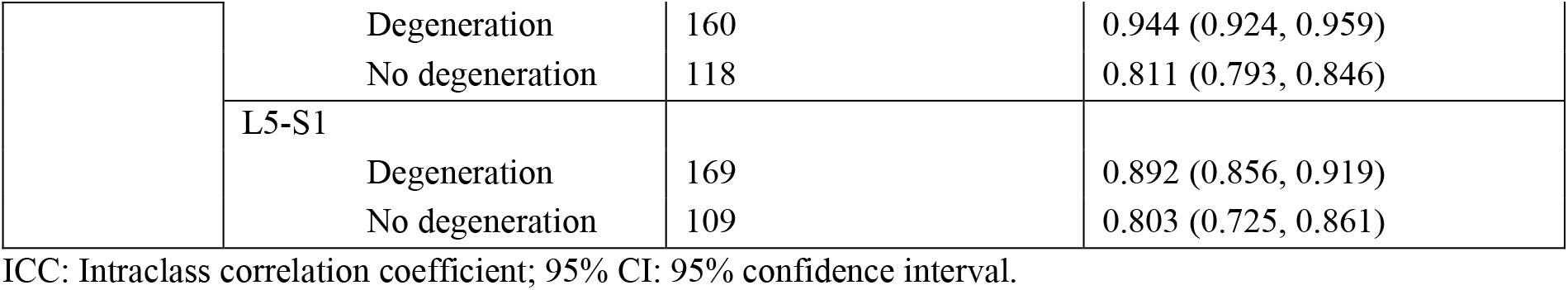
Inter-rater measures’ reliability results of disc height index (DHI) on all segmental levels in degeneration and no degeneration group

**ESM_2_Table 2.**
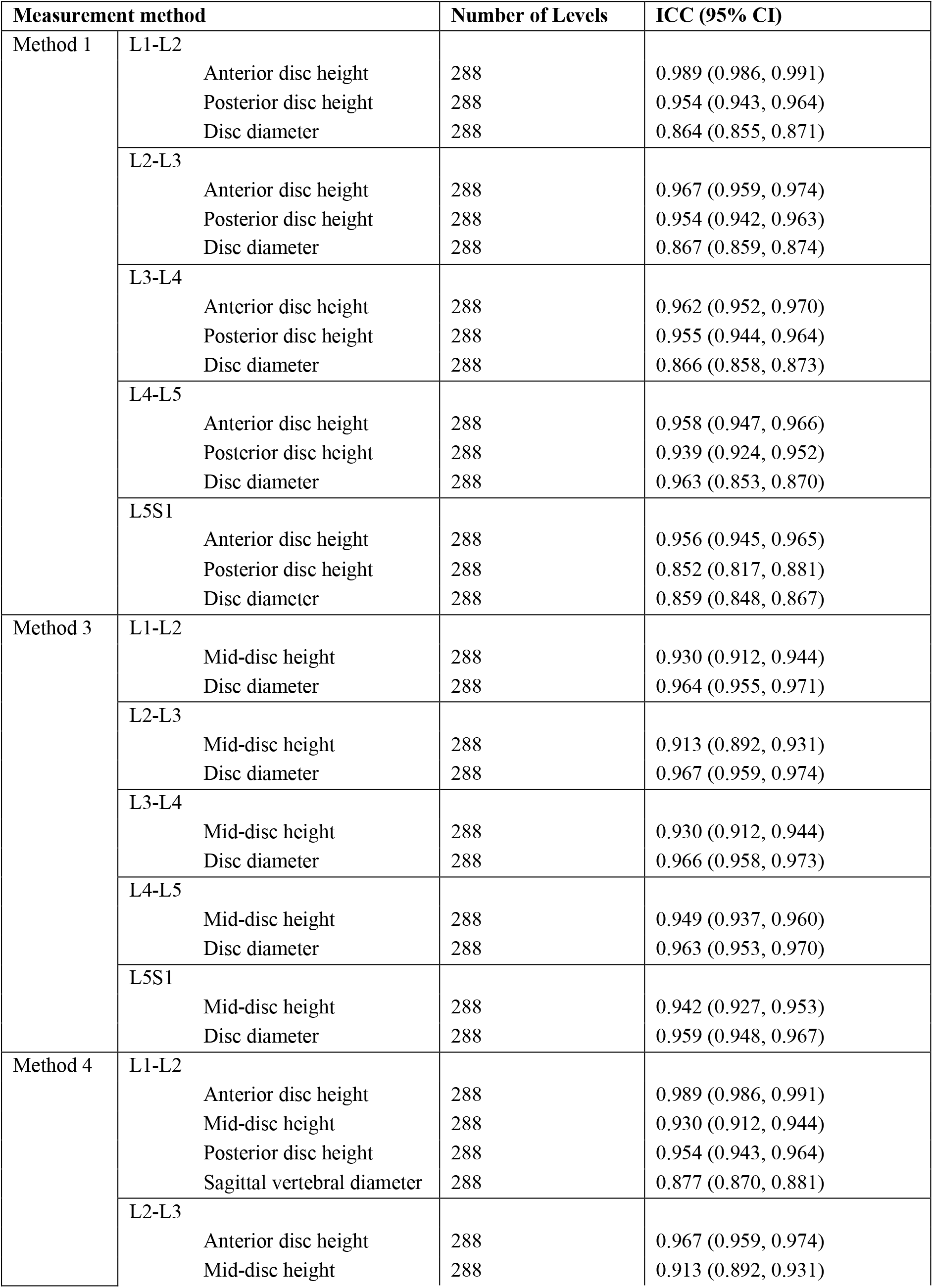
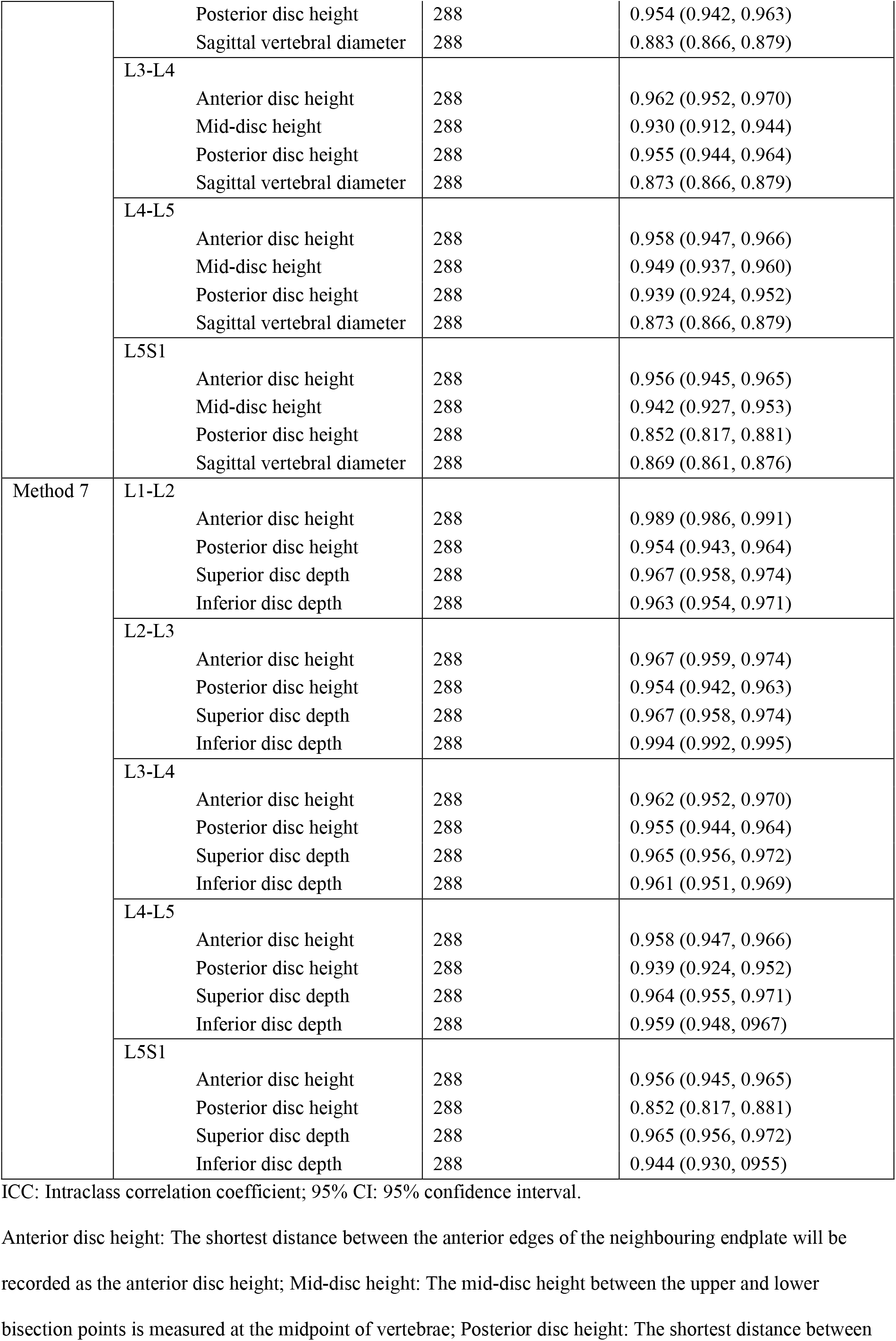
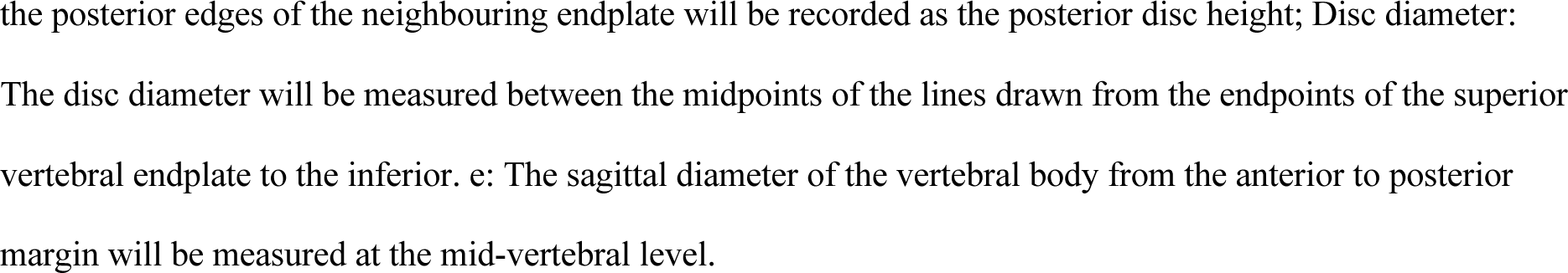
Inter-rater measures’ reliability results of related lines to each measurement method (method could be used on all segment levels and ICCs are good-to-excellent) on disc height index (DHI) of all segmental levels

**ESM_2_Table 3.** Potential factors for the outliers (out of the 95% Limits of Agreement (LOA) on the Bland and Altman plot)

| Method | Level | Number of outliers | Potential factor |
| --- | --- | --- | --- |
| Method 1 | L1-L2 | 12 | 9 outliers of nucleus pulposus degeneration, 3 outliers of disc herniation |
|  | L2-L3 | 19 | 15 outliers of nucleus pulposus degeneration, 4 outliers of disc herniation |
|  | L3-L4 | 10 | 8 outliers of nucleus pulposus degeneration, 1 outlier of disc herniation, 1 normal |
|  | L4-L5 | 2 | 2 outliers of nucleus pulposus degeneration |
|  | L5-S1 | 14 | 7 outliers of nucleus pulposus degeneration, 6 outliers of disc herniation, 1 normal |
|  | All | 57 | 41 outliers of nucleus pulposus degeneration, 14 outliers of disc herniation, 2 normal |
| Method 2 | L1-L2 | 15 | 9 outliers of nucleus pulposus degeneration, 5 outliers of disc herniation, 1 normal |
|  | L2-L3 | 15 | 10 outliers of nucleus pulposus degeneration, 5 outliers of disc herniation |
|  | L3-L4 | 13 | 10 outliers of nucleus pulposus degeneration, 3 outliers of disc herniation |
|  | L4-L5 | 10 | 8 outliers of nucleus pulposus degeneration, 2 outliers of disc herniation |
|  | L5-S1 | 12 | 6 outliers of nucleus pulposus degeneration, 5 outliers of disc herniation, 1 normal |
|  | All | 65 | 43 outliers of nucleus pulposus degeneration, 20 outliers of disc herniation, 2 normal |
| Method 3 | L1-L2 | 37 | 21 outliers of nucleus pulposus degeneration, 15 outliers of disc herniation, 1 normal |
|  | L2-L3 | 26 | 20 outliers of nucleus pulposus degeneration, 6 outliers of disc herniation |
|  | L3-L4 | 21 | 16 outliers of nucleus pulposus degeneration, 3 outliers of disc herniation, 2 normal |
|  | L4-L5 | 45 | 38 outliers of nucleus pulposus degeneration, 13 outliers of disc herniation, 4 normal |
|  | L5-S1 | 42 | 24 outliers of nucleus pulposus degeneration, 17 outliers of disc herniation, 1 normal |
|  | All | 171 | 109 outliers of nucleus pulposus degeneration, 54 outliers of disc herniation, 8 normal |
| Method 4 | L1-L2 | 57 | 34 outliers of nucleus pulposus degeneration, 17 outliers of disc herniation, 6 normal |
|  | L2-L3 | 5 | 4 outliers of nucleus pulposus degeneration, 1 outlier of disc herniation |
|  | L3-L4 | 42 | 31 outliers of nucleus pulposus degeneration, 7 outliers of disc herniation, 4 normal |
|  | L4-L5 | 44 | 29 outliers of nucleus pulposus degeneration, 11 outliers of disc herniation, 4 normal |
|  | L5-S1 | 34 | 18 outliers of nucleus pulposus degeneration, 15 outliers of disc herniation, 1 normal |
|  | All | 182 | 116 outliers of nucleus pulposus degeneration, 51 outliers of disc herniation, 15 normal |
| Method 5 | L1-L2 | 1 | 1 outlier of nucleus pulposus degeneration |
|  | L2-L3 | 12 | 6 outliers of nucleus pulposus degeneration, 6 outliers of disc herniation |
|  | L3-L4 | 10 | 6 outliers of nucleus pulposus degeneration, 4 outliers of disc herniation |
|  | L4-L5 | 14 | 8 outliers of nucleus pulposus degeneration, 6 outliers of disc herniation |
|  | All | 37 | 21 outliers of nucleus pulposus degeneration, 16 outliers of disc herniation |
| Method 6 | L1-L2 | 11 | 5 outliers of nucleus pulposus degeneration, 6 outliers of disc herniation |
|  | L2-L3 | 10 | 7 outliers of nucleus pulposus degeneration, 3 outliers of disc herniation |
|  | L3-L4 | 13 | 9 outliers of nucleus pulposus degeneration, 4 outliers of disc herniation |
|  | L4-L5 | 8 | 6 outliers of nucleus pulposus degeneration, 2 outliers of disc herniation |
|  | All | 42 | 27 outliers of nucleus pulposus degeneration, 15 outliers of disc herniation |
| Method 7 | L1-L2 | 6 | 5 outliers of nucleus pulposus degeneration, 1 outlier of disc herniation |
|  | L2-L3 | 17 | 14 outliers of nucleus pulposus degeneration, 3 outliers of disc herniation |
|  | L3-L4 | 17 | 13 outliers of nucleus pulposus degeneration, 3 outliers of disc herniation, 1 normal |
|  | L4-L5 | 1 | 1 outlier of nucleus pulposus degeneration |
|  | L5-S1 | 14 | 4 outliers of nucleus pulposus degeneration, 9 outliers of disc herniation, 1 normal |
|  | All | 55 | 37 outliers of nucleus pulposus degeneration, 16 outliers of disc herniation, 2 normal |

